# Dysphagia and associated pneumonia in stroke patients from India

**DOI:** 10.1101/2020.02.26.20028159

**Authors:** Rahul Krishnamurthy, Priya Karimuddanahally Premkumar, Radish Kumar Balasubramanium

## Abstract

**Background:** India has high incidence (116-163 per 100,000) of stroke compared to western countries. Stroke is reported to be the fourth leading cause of death and fifth leading cause of disability in India. Dysphagia is seen approximately in half of the stroke patients and if unidentified may result in pulmonary complications such as aspiration pneumonia. However, there is no estimate of post stroke dysphagia and associated pneumonia in India.

**Method:** Using the PRISMA methodology, a systematic search for recent literature on dysphagia in stroke was carried out across all the major databases. Two authors independently screened the titles and the abstracts, and those selected articles were assessed for quality using the GRADE approach. Comparisons were made of reported dysphagia and pneumonia frequencies, the relative risks of developing pneumonia were calculated. Data on duration of hospital stay and rates of mortality were also extracted.

**Results:** We identified 86 citations, out of which only four articles were deemed eligible for critical analyses and data extraction. A high incidence (11.1% - 87.5%) of dysphagia was observed among stroke patients. Only two studies reported on incidence of pneumonia (22.8% - 32%); only one study stratified patients by both dysphagia and pneumonia. A relative risk of 5.82 (95% CI 4.6, 7.2) was found for pneumonia in patients with stroke and dysphagia. Data on length of hospital stay and rates of mortality secondary to aspiration pneumonia are also presented.

**Conclusion:** Despite high incidence of dysphagia and associated pneumonia the methodological quality of studies are low. There is a dire need for methodologically sound studies to accurate determine the incidence of dysphagia and its impact on stroke patients in India.

---

The incidence of stroke in India is estimated to be around 116-163 per 100,000 population.^1^ A recent report from the Indian Council of Medical Research (ICMR)^2^ titled ‘India: Health of the nation’s states,’ suggests stroke to be the fourth leading cause of death and fifth leading cause of disability adjusted life years (DALY) in India. As an initiative to promote better stroke care, the government of India through the National Programme for Prevention and Control of Cancer, Diabetes, Cardiovascular Diseases & Stroke (NPPCDCS)^3^ has developed guidelines for prevention and management of stroke. These guidelines align with those of North America (United States^4^ and Canada^5^) and the United Kingdom^6^. Similar to its global counterparts, the Indian guidelines recommend dysphagia screening for all conscious patients. However, organized rehabilitation services in the country are limited and few health care professionals have specialized training in stroke. Furthermore, specialized stroke centers are predominantly limited to urban centers only. Due to these reasons, the majority of stroke survivors continue to live with disabilities and suffer post stroke complications.

Dysphagia is seen approximately in half of all the stroke patients^7,8^ and if unidentified may result in pulmonary complications such as aspiration pneumonia.^7,9,10^ Along with these life-threatening complications, dysphagia is also associated with substantial healthcare costs. In the western scenario, a study by Altman et al^11^ report that the annual attributable cost of dysphagia is around $547 million. Patel et al.^12^ analyzed the healthcare cost of dysphagia in the United States using the National Inpatient Sample Database of the Agency for Healthcare Research and Quality (AQHR) between periods of 2009 to 2013. They report that the estimated additional cost of dysphagia for the study period was $16.8 billion. The period of hospitalization was found to be 8.8 days for those individuals with dysphagia versus 5 days for those without dysphagia. Even though the exact healthcare costs of dysphagia in India in not known, the burden of dysphagia is significant as the costs of ongoing rehabilitation and long-term care are largely overseen by family members, which burdens the family further.

A systematic review by Kishore et al^13^ report that the rate of pneumonia among stroke patients is around 14% globally and the rate increases by eight times among individuals with dysphagia. Another study by Pacheco□Castilho et al^14^ aimed to estimate dysphagia and pneumonia incidence among stroke patient in Brazil. The findings of the study reveal that the frequency of dysphagia among stroke patients was around 59% to 76%, and the relative risk for pneumonia in patients with stroke and dysphagia was around 8.4. To date there have been no collective reports on estimate of dysphagia incidence among stroke patients in India. For a country like India, this epidemiological data is crucial to allocate limited national resources, maximize stroke care, set future benchmarks for optimal intervention, and influence national health policies. In an attempt to address the existing knowledge gap, the present systematic review is undertaken. The present study aimed to systematically review the existing literature on patients with stroke, synthesize data on estimates of dysphagia incidence and associated aspiration pneumonia risks in India.

## Method

### Objectives of the study

1. To identify the reported frequency of oropharyngeal dysphagia among stroke patients in India.
2. To identify frequency of associated aspiration pneumonia risk among stroke patients with and without oropharyngeal dysphagia in India.
3. To identify national trends in dysphagia assessment.

### Operational definitions

The following operational definitions, as defined by Pacheco□Castilho et al^14^ were adopted in the present study.

- *Oropharyngeal dysphagia*: defined as any physiological impairment affecting the oral, pharyngeal and/or upper esophageal phases of swallowing.
- *Pneumonia*: defined as any infection in one or both of the lungs (if pneumonia was reported, the criteria by which it was defined had to be declared).
- *Stroke*: defined as any confirmed diagnosis by medical and/or imaging exams and treated in acute, rehabilitation, or chronic facilities (public and/or private) and regardless of stroke type or location.

### Information source and database search

The present systematic review is consistent with the PRISMA^15^ statement. Systematic searches were carried out between October 2019 and December 2019 in the following databases: PubMed, CINAHL, ProQuest, Ovid, Scopus, and Cochrane. The reference lists of related articles were also screened for eligible papers. A search strategy (see appendix for full search strategies) for each of the database was developed using the keywords “deglutition, deglutition disorders, swallowing, swallowing disorders, swallowing problems, swallowing difficulties, oropharyngeal, dysphagia, stroke, cerebrovascular diseases, and India” along with Boolean operators, such as “AND” and “OR.”

### Study selection

The present review adopted a three-phase selection process for the final inclusion of studies. The studies obtained from the search of all the above-mentioned databases were compiled using the Mendeley desktop reference management system. The duplicates were removed, and the authors of the present study independently screened the titles. Inappropriate study titles were excluded from further screening. Review articles, case reports, editorials were excluded; only those articles published in peer-reviewed journals and dissertations in the English language were considered for further review.

The second phase involved the screening of those abstract titles selected from the first phase. Both the authors independently reviewed those titles and any disagreements that arose were discussed to arrive at a consensus. Those abstract titles selected at the end of phase two were considered for full article review. In addition, the reference lists of the full-length articles were manually searched to identify any potential articles that could have been missed.

### Analysis of risk of bias

The Cochrane review methodology^16^ was used to assess each included article for the risk of bias. Each article was assessed in five main domains: Selection bias, performance bias, detection bias, attrition bias, and reporting bias. Initially, the first author assessed and rated all the finalized articles. Following this, the second author checked the rating accuracy for risk data. Any disagreements that arose were mutually resolved by consensus.

### Level of evidence analysis and Methodological quality appraisal

The Grades of recommendation, assessment, development, and evaluation working group (GRADE) approach^16^ was used to analyze the level of evidence of each included article. This approach determines the quality of the study based on the methodology employed in it. Study designs such as randomized control trials (RCTs) are highly rated, observational studies receive a moderate grade, and a low rating is given to case studies. The authors, based on literature developed a methodological quality appraisal tool (shown in table 1) suitable for the present systematic review. The scoring percentage was calculated by dividing the score obtained divided by total and multiplied by 100 for each study. The scoring percentage was used to categorize the studies included: 0–33.9% as weak, 34%–66.9% as moderate, and above 67% as strong.

**Table 1:** Methodological quality appraisal tool for included studies.

| No. | Question |
| --- | --- |
| 1. | Was the aim or objective(s) of the study clearly mentioned |
| 2. | Was the participant details clearly mentioned and defined |
| 3. | Was the sample size justification provided |
| 4. | Are the findings for specified objectives clearly mentioned |
| 5. | Are the procedures, instruments used in the study specified and explained |
| 6. | Was the method of assessment reproducible? |
| 7. | Was the method of assessment validated? |
| 8. | Were all the patients included in the analysis? |
| 9. | Were all the pre-specified outcomes reported? |
| 10. | Were all the parameters mentioned in the objectives explained in results? |

*Note:* Rating 1 = yes, 0 = no / not reported

## Results

### Literature retrieval

The PRISMA chart of the systematic review procedure has been illustrated in figure 1.

**Figure 1:**
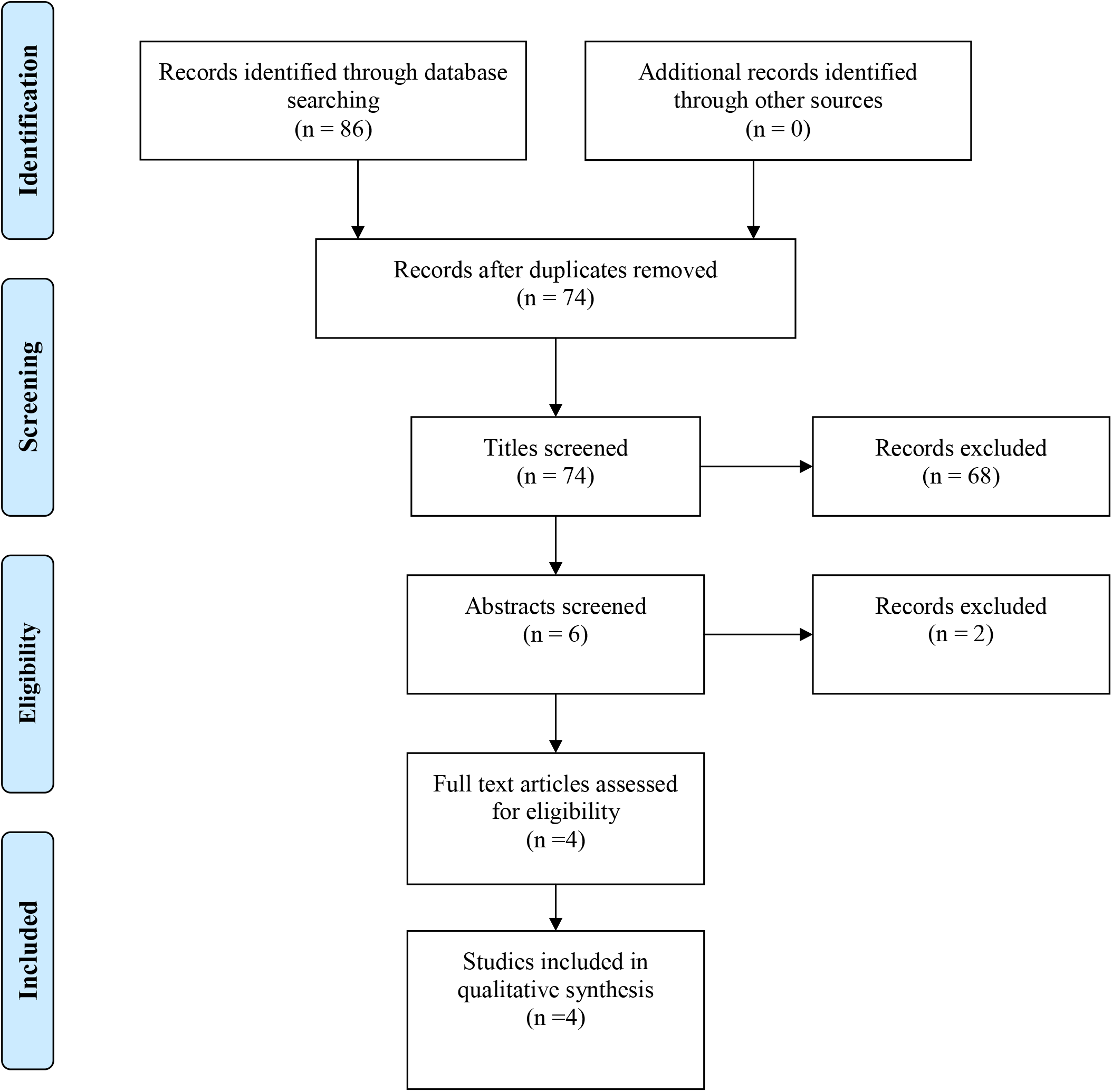
PRISMA chart of the systematic review procedure.

We identified 86 citations across all the databases. After an initial screening, 12 duplicates were obtained and excluded. The remaining 74 titles were screened for eligibility, of which 6 articles studies were accepted for abstract review. Of these articles, two did not meet our inclusion criteria and only four were found to be suitable for inclusion.

### Study characteristics

Methodological appraisal of the included studies was carried out using a custom developed tool (shown in table 1), which was based on the literature review. The scoring for each of the included studies is mentioned in table 2.

**Table 2:** Methodological quality rating for the studies included

| No. | Study | Q1 | Q2 | Q3 | Q4 | Q5 | Q6 | Q7 | Q8 | Q9 | Q10 | Score |
| --- | --- | --- | --- | --- | --- | --- | --- | --- | --- | --- | --- | --- |
| 1. | Sreeraj et al<sup>17</sup> | 1 | 0 | 1 | 1 | 0 | 0 | 0 | 1 | 0 | 0 | 40% |
| 2. | Rai et al<sup>18</sup> | 1 | 1 | 0 | 1 | 1 | 0 | 0 | 0 | 1 | 1 | 60% |
| 3. | Sundar et al<sup>19</sup> | 1 | 0 | 0 | 0 | 1 | 1 | 1 | 0 | 0 | 1 | 60% |
| 4. | Sebastian et al<sup>20</sup> | 1 | 0 | 0 | 1 | 1 | 1 | 0 | 0 | 1 | 1 | 60% |

*Note: Rating 1= yes; 0= no/not reported*

Based on the score %, it was observed that all the studies considered for the final review have moderate quality. Based on the GRADE approach, the included studies have a low level of evidence. The study characteristics of the included articles have been summarized in table 3.

### Age, sex, and type of stroke

Across the included studies, the number of participants sampled per study ranged from 16 participants^20^ to 486 participants^17^. Age related data was only reported by Sreeraj et al^17^ (18 to 90 years) study, and the other studies^18-20^ did not report any data with respect to age of the participants. Sex distribution of the participants was not clearly described in any of the four studies.^17-20^ All the four studies^17-20^ included in the final review consisted of both ischemic and hemorrhagic stroke.

### Trends in dysphagia assessment

Across all the studies, clinical assessment was used for the diagnosis of dysphagia. The one exception was Sundar et al^19^ study which used pulse oximetry and clinical assessment. It was noted that none of the studies utilized standardized clinical assessment protocols/tests. Mostly, the assessment was carried out by a resident physician and not by a speech language pathologist. The food consistency used to assess swallow varied with most studies. The study by Rai et al^18^ used ‘Khichri/Kheer’ (based on the North Indian diet), or Payasam (based on the South Indian diet), which they describe as equivalents of the western puree thick consistency. Sundar et al^19^ used liquid thin consistency (50 ml water) only. The other two studies^17,20^ did not describe the consistency used to assess swallowing. The co-occurrence of dysphagia and pneumonia was assessed only in two studies.^17,19^ None of the studies operationally defined pneumonia.

### Frequency of dysphagia and associated pneumonia

All the four studies utilized clinical assessments to identify dysphagia. The frequency of identified dysphagia ranged from 11.1%^18^ to 87.5%.^20^ However, only two studies^17,19^ reported occurrence of associated pneumonia. In Sreeraj et al^17^ study, aspiration pneumonia was reported in 91 (22.8%) patients among 154 patients identified with dysphagia. Sundar et al^19^ reported that 16 (32%) patients developed aspiration pneumonia among 21 patients identified with dysphagia. This data was used to calculate relative risk for pneumonia in patients with stroke and dysphagia versus those patients without dysphagia. We found a relative risk of 5.82 (95% CI 4.6, 7.2) for pneumonia in patients with stroke and dysphagia versus the same patients with stroke and no dysphagia.

### Dysphagia, associated pneumonia, duration of hospital stay, and mortality

Sundar et al^19^ reported that the average length of hospital stay for those patients with dysphagia was around 10.8 days versus 6.5 days for those without dysphagia. Even though Sreeraj et al^17^ do not report exact duration of hospital stay, they report an odds ratio of 1.9 (CI 1.2, 2.9) for those individuals with dysphagia as compared to those without dysphagia.

Rates of mortality associated with aspiration pneumonia secondary to dysphagia was reported only by two studies^17,19^. Sreeraj et al^17^ report an odds ratio of 10.99 (6.3, 19.1) for mortality among those individual with aspiration pneumonia secondary to dysphagia compared to those without aspiration pneumonia. In Sundar et al^19^ study it was found that five out of 16 patients who developed aspiration pneumonia expired.

## Discussion

The present study is an initial attempt to review existing data on dysphagia and associated pneumonia among stroke patients in India. We identified very few studies that met our inclusion criteria; however, we found that dysphagia was a common consequence among patients with stroke in India. The findings of the present study highlight the importance of implementing uniform strategies to identify and manage dysphagia in stroke population to prevent further complications. Unfortunately, the quality of the available literature is low with potential risks for bias across several categories. We believe that these biases may limit both the internal and the external validity of the frequency estimates made in the present study.

The first objective of the study was to investigate the national trends assessment of oropharyngeal dysphagia in stroke population. Our results showed that the national trends in dysphagia assessment did not adequately capture the entirety of oropharyngeal swallow physiology. Mostly, clinical swallowing examination alone was used to diagnose dysphagia. It was also observed that the clinical swallowing examination was majorly carried out utilizing only the clinical signs of aspiration, whereas no standardized tests with established validity and reliability were used. The use of non standardized clinical assessment tools may have likely underestimated the presence of dysphagia and events of silent aspiration may have been missed significantly. None of the studies reviewed in the present article had utilized instrumental examination. This trend is likely a reflection of the current limitation in most of Indian medical services for assessment of dysphagia.

The estimates of dysphagia among individuals with stroke in India (between 11.6 and 87.5%) is higher than the estimates reported in the western studies. A systematic review of studies from developed countries by Martino et al^7^ report a much lower estimates between 51 – 55%. Similar trends are observed in studies of cohorts from developed countries such as Spain (47%),^23^ Canada (45%),^24^ and Italy (50%).^9^ The estimates obtained in this systematic review is also higher than some studies in emerging countries such as South Africa (53%),^25^ and Brazil (59 – 76%).^14^

The rate of pneumonia in India was found to be between 22.8 to 32%, which is twice as higher than the global pneumonia rate (15%), and also was higher when compared to other emerging countries such as Brazil (15%),^26^ and Chile (23%).^27^ However, this increased pneumonia risk in India could only be derived from two studies.^17,19^ Using the data from two studies,^17,19^ a relative risk for pneumonia in stroke was found to be 5.82 (95% CI 4.6, 7.2). These estimates is higher than that of western countries [3.2 (95% CI 2.1, 4.9)] as reported by Martino et al,^7^ but was lower than that of Brazil [8.4 (95% CI 2.1, 34.4)] as reported by Pacheco-Castilho et al.^14^ Furthermore, pneumonia was not operationally defined in any of the two studies, thus limiting its validity.

### Limitations

As with all systematic reviews, the findings from our study are limited by the quality of the original studies included for the review. Especially, the studies included did not provide crucial information known to affect dysphagia presence, such as: stroke type, multiple stroke events, site of lesion, severity of stroke, time of assessment, consistency and volume of bolus used for clinical assessment of swallowing, and the definitions adopted for dysphagia and pneumonia. All the studies included in the review presented with a potential detection bias, as dysphagia was diagnosed subjectively using clinical evaluation tools without adequate psychometric validation. These methodological inconsistencies may have influenced in overestimating or underestimating frequency of dysphagia and associated pneumonia.

### Conclusion

The current systematic review is an attempt to estimate incidence of dysphagia and associated pneumonia among stroke patients in India. The data from this systematic review is crucial for health care professionals to prioritize early detection of dysphagia and its management. Findings from our study reveal that quantity and quality of studies focused on dysphagia and associated pneumonia among stroke patients in India is low. Methodologically strong studies are needed to obtain better understanding of dysphagia and its concomitant pneumonia risk among stroke patients in India. These studies would be crucial to appropriately inform future updates on optimization of dysphagia care among stroke patients in India.

## Data Availability

Nil

## Acknowledgements

**Nil**

